# Homologous and Heterologous Vaccine Boost Strategies for Humoral and Cellular Immunologic Coverage of the SARS-CoV-2 Omicron Variant

**DOI:** 10.1101/2021.12.02.21267198

**Authors:** C. Sabrina Tan, Ai-ris Y. Collier, Jinyan Liu, Jingyou Yu, Abishek Chandrashekar, Katherine McMahan, Huahua Wan, Xuan He, Catherine Jacob-Dolan, Daniel Sellers, John D. Ventura, Yannic Bartsch, Blake M. Hauser, Jennifer E. Munt, Melissa Mattocks, Kathryn E. Stephenson, Samuel J. Vidal, Kate Jaegle, Marjorie Rowe, Rachel Hemond, Lorraine Bermudez Rivera, Tochi Anioke, Julia Barrett, Benjamin Chung, Sarah Gardner, Makda S. Gebre, Nicole Hachmann, Michelle Lifton, Jessica Miller, Felix Nampanya, Olivia Powers, Michaela Sciacca, Mazuba Siamatu, Nehalee Surve, Lisa H. Tostanoski, Haley VanWyk, Cindy Wu, Ralph S. Baric, Aaron G. Schmidt, Galit Alter, Dan H. Barouch

## Abstract

The rapid spread of the highly mutated SARS-CoV-2 Omicron variant has raised substantial concerns about the protective efficacy of currently available vaccines. We assessed Omicron-specific humoral and cellular immune responses in 65 individuals who were vaccinated with two immunizations of BNT162b2 and were boosted after at least 6 months with either Ad26.COV2.S (Johnson & Johnson; N=41) or BNT162b2 (Pfizer; N=24) (Table S1).

---

The rapid spread of the highly mutated SARS-CoV-2 Omicron variant has raised substantial concerns about the protective efficacy of currently available vaccines. We assessed Omicron-specific humoral and cellular immune responses in 65 individuals who were vaccinated with two immunizations of BNT162b2 and were boosted after at least 6 months with either Ad26.COV2.S^1^ (Johnson & Johnson; N=41) or BNT162b2^2^ (Pfizer; N=24) (**Table S1**).

Six months following BNT162b2 vaccination, pseudovirus neutralizing antibody responses were detectable but low to the WA1/2020, Delta, and Beta variants^3,4^ but were largely undetectable to the Omicron variant, indicating substantial immune escape (**Fig. 1A**). Ad26.COV2.S boosted median Omicron-specific neutralizing antibody titers from 21 at week 0 prior to the boost to 591 at week 2 following the boost, and these titers further increased to 859 at week 4, reflecting a 41-fold increase compared with pre-boost levels (**Fig. 1A, Table S2**). BNT162b2 boosted median Omicron-specific neutralizing antibody titers from 20 at week 0 to 1,018 at week 2, but these titers declined to 345 at week 4, reflecting a 17-fold increase compared with pre-boost levels (**Fig. 1A, Table S2**). Similar boost kinetics were observed by live virus neutralization assays to the WA1/2020 and Beta variants (**Fig. S1**).

**Figure 1.**
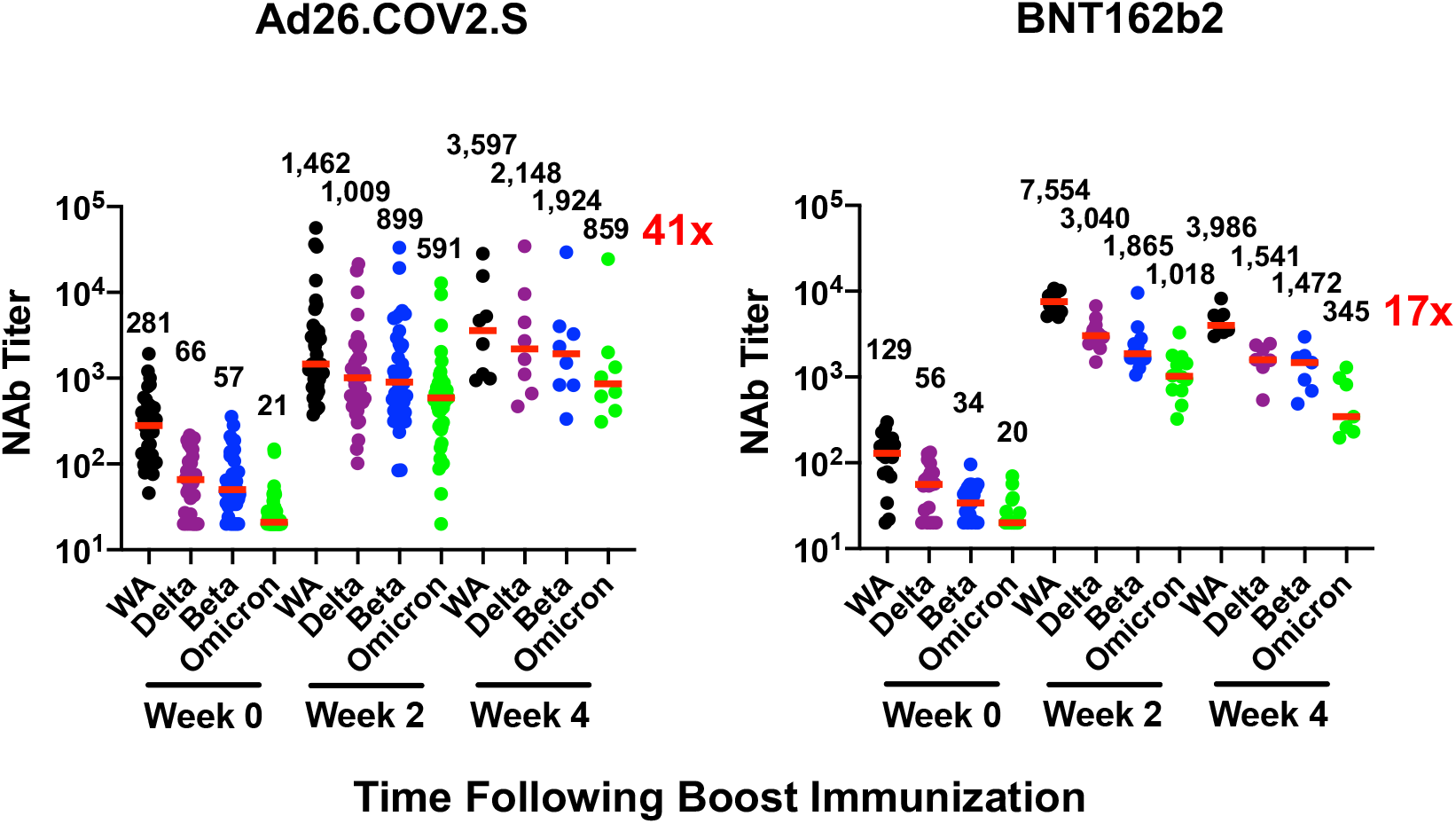
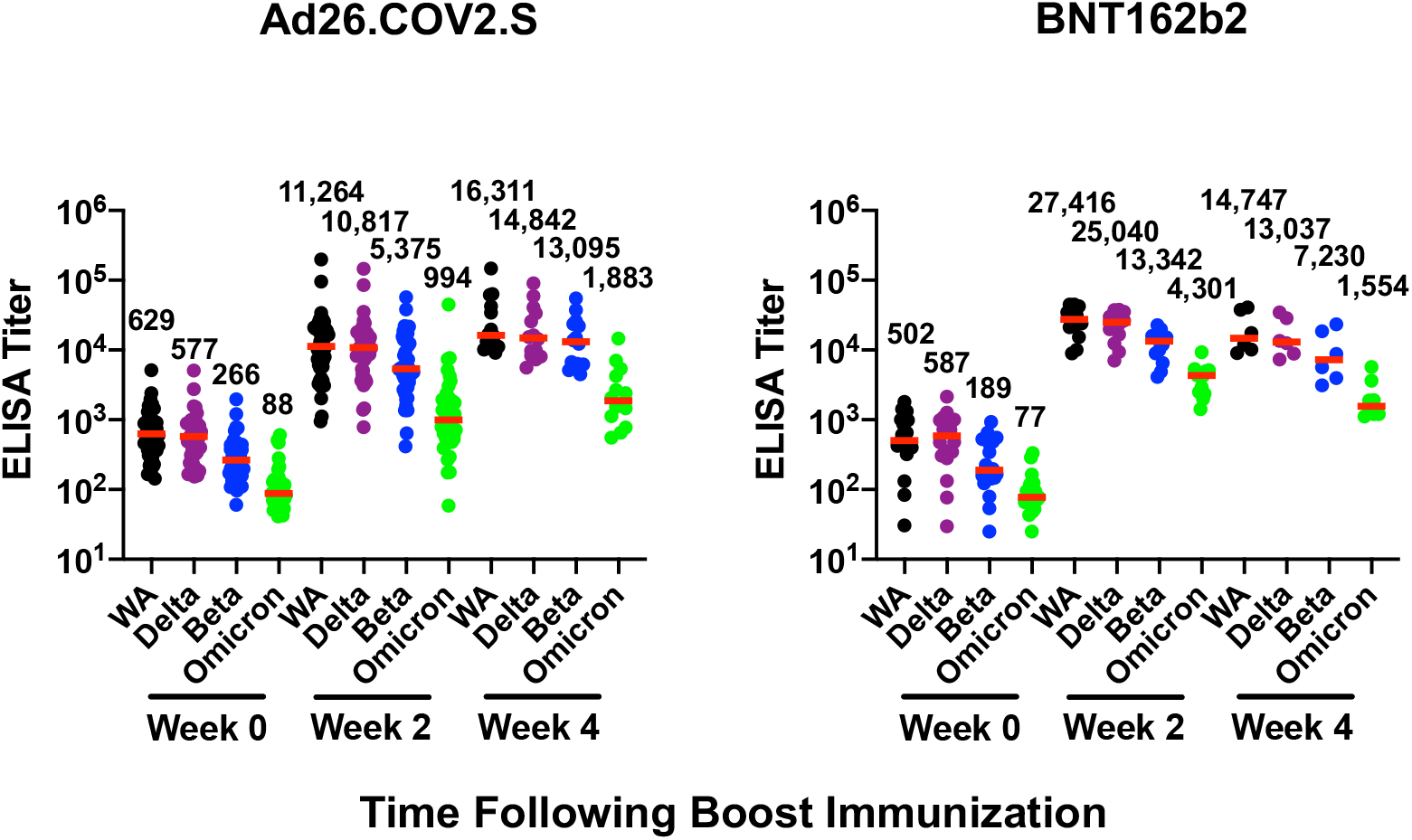
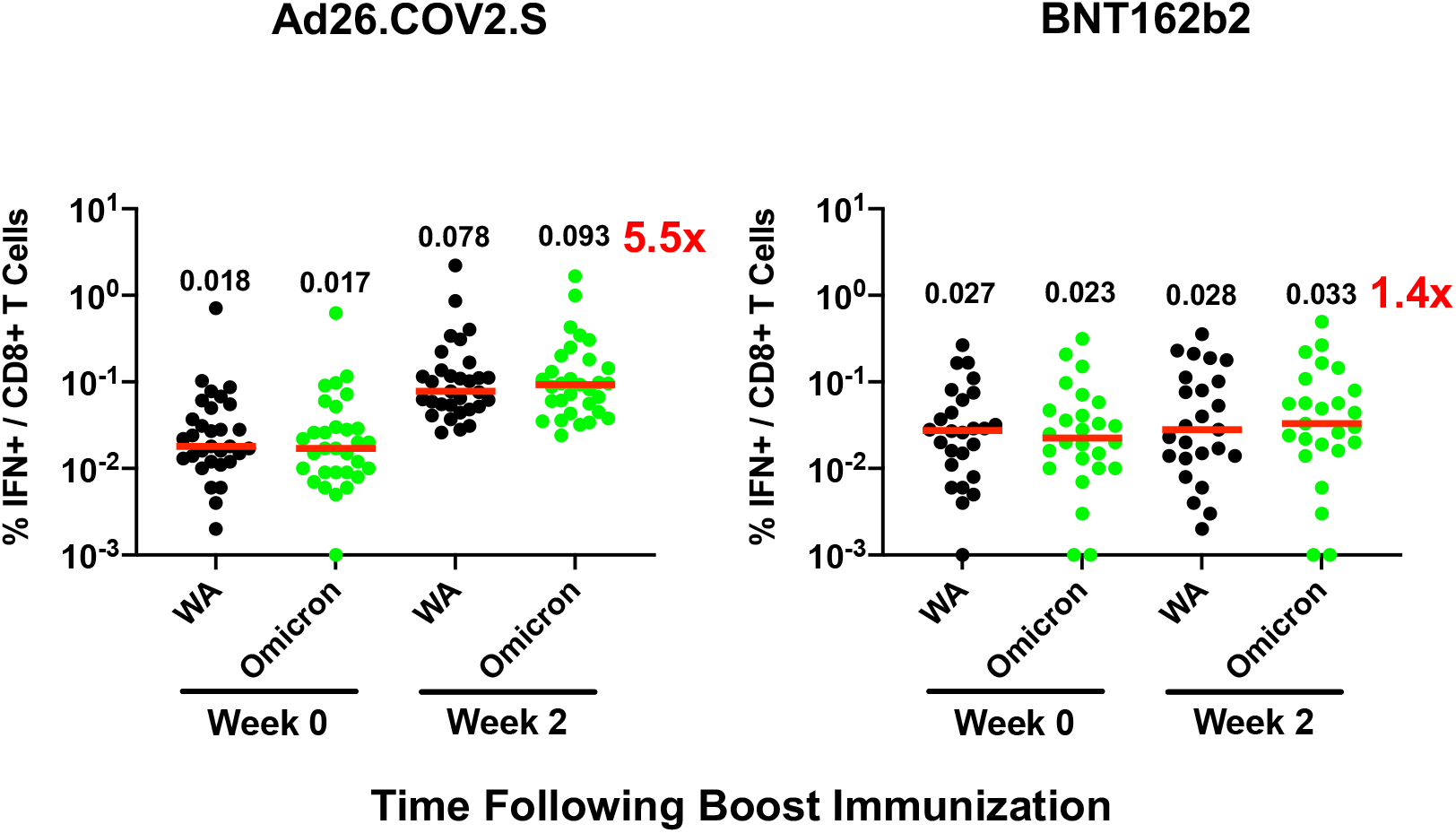
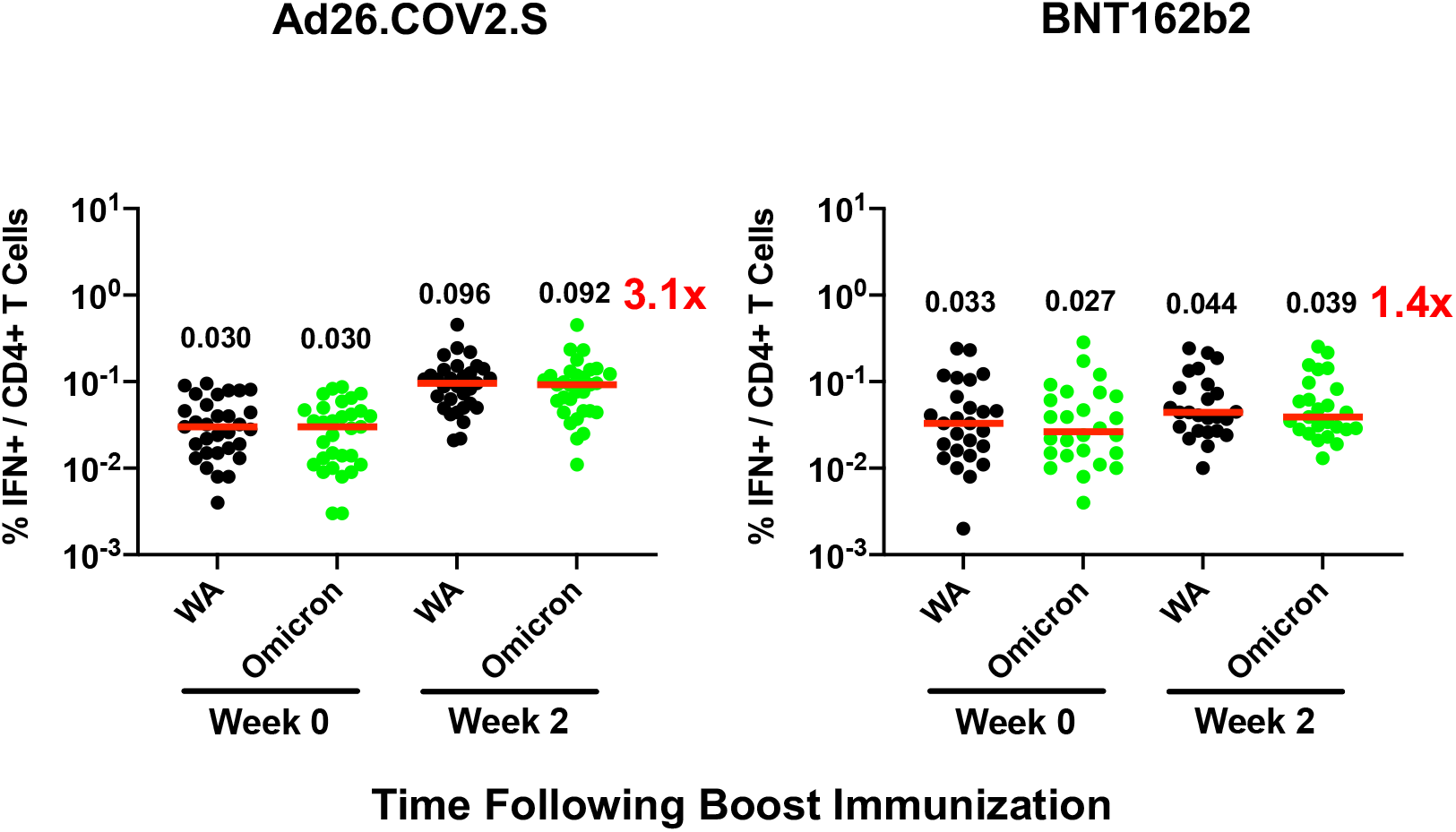
Humoral and cellular immune responses following Ad26.COV2.S and BNT162b2 boosting. Immune responses at weeks 0, 2, and 4 following boosting BNT162b2 vaccinated individuals with Ad26.COV2.S or BNT162b2. **A**. Neutralizing antibody (NAb) titers by a luciferase-based pseudovirus neutralization assay. **B**. Receptor binding domain (RBD)-specific binding antibody titers by ELISA. Pooled peptide Spike-specific IFN-*γ* CD8+ T cell responses (**C**) and CD4+ T cell responses (**D**) by intracellular cytokine staining assays. Responses were measured against the SARS-CoV-2 WA1/2020, B.1.617.2 (Delta), B.1.351 (Beta), and B.1.1.529 (Omicron) variants. Medians (red bars) are depicted and numerically shown. A subset of samples were available at week 4.

Ad26.COV2.S boosted median Omicron-specific binding antibody titers by ELISA from 88 at week 0 to 994 at week 2 and 1,883 at week 4 (**Fig. 1B**). BNT162b2 boosted median Omicron-specific binding antibody titers from 77 at week 0 to 4,301 at week 2 and 1,554 at week 4 (**Fig. 1B**). Similar boost kinetics were observed by electrochemiluminescence binding antibody assays (**Fig. S2**) and IgG isotype-specific binding antibody assays (**Fig. S3**). These data show that both heterologous and homologous boosting strategies led to large increases in Omicron-specific antibody responses. Ad26.COV2.S boosting led to peak antibody titers at week 4 or later, whereas BNT162b2 boosting led to peak antibody titers at week 2 that declined by week 4.

Although antibody responses were substantially reduced to Omicron compared with WA1/2020, CD8+ and CD4+ T cell responses were similar for Omicron and WA1/2020, suggesting substantial cellular immune cross-reactivity (**Fig. 1C, 1D**). Ad26.COV2.S boosted median Omicron-specific CD8+ T cells by 5.5-fold from 0.017% to 0.093% (**Fig. 1C**) and Omicron-specific CD4+ T cells by 3.1-fold from 0.030% to 0.092% (**Fig. 1D**). Ad26.COV2.S also increased Omicron-specific central and effector memory CD8+ and CD4+ T cell responses (**Fig. S4, S5**). BNT162b2 increased Omicron-specific CD8+ and CD4+ T cells by 1.4-fold (**Fig. 1C, 1D**). Ad26.COV.2 and BNT162b2 also increased WA1/2020-specific memory B cell responses (**Fig. S6, S7**).

Both heterologous “mix-and-match” Ad26.COV2.S boosting and homologous BNT162b2 boosting increased Omicron-specific humoral and cellular immune responses in individuals who were vaccinated at least 6 months previously with BNT162b2, although these boosting strategies showed different immune kinetics (**Fig. S8**). Omicron-specific antibody responses were markedly reduced compared with WA1/2020-specific antibody responses, but Omicron-specific T cell responses were highly cross-reactive, and preclinical studies have shown that CD8+ T cells are critical for protection against SARS-CoV-2 disease when antibody responses are suboptimal^5^. The durability and protective efficacy of heterologous and homologous boost vaccine strategies for the SARS-CoV-2 Omicron variant remain to be determined.

## Data Availability

All data produced in the present work are contained in the manuscript

## Data Availability

The datasets generated during and/or analysed during the current study and the analysis codes used to produce the figures and tables are available in the Open Science Framework repository, https://doi.org/10.17605/OSF.IO/HQNKR.

## Data sharing

C.S.T, A.Y.C., and D.H.B. had full access to all the data in the study and take responsibility for the integrity of the data and the accuracy of the data analysis. All data are available in the manuscript or the supplementary material. Correspondence and requests for materials should be addressed to D.H.B.

## Author Contributions

CST, AYC, DHB: conceptualization, formal analysis, resources, investigation, data curation, writing-original draft, writing-review & editing, visualization, supervision

JL, JY, AC, KAM, HW, XH, CJD, DS, JV, RSB, AGS, GA: investigation, methodology, data curation, writing-review & editing, supervision

YB, BMH, JEM, MM, KES, SJV, KJ, JLC, MR, RH, LBR, TA, JB, BC, SG, MG, NH, ML, JM, FM, OP, MSc, MSi, NS, LHT, HV, CW: resources, methodology, writing-review & editing

## Acknowledgements

The authors thank the study participants, the Center for Virology and Vaccine Research, and the Department of Obstetrics and Gynecology for enrollment, collection, and processing samples for the BIDMC COVID-19 Biorepository. We thank MesoScale Discovery for providing the kits for the ECLA multiplexing kits used in this study.

## Supplementary Methods

### Study population

A specimen biorepository at Beth Israel Deaconess Medical Center (BIDMC) obtained samples from individuals who received the BNT162b2 vaccine. The BIDMC institutional review board approved this study (2020P000361). Participants either continued follow-up in the biorepository and were boosted with BNT162b2 clinically or were enrolled in the COV2008 study (NCT04999111) and were boosted with Ad26.COV2.S. An additional 16 participants who were not enrolled in the BIDMC biorepository were also enrolled in the COV2008 study. The Advarra institutional review board approved the COV2008 study (SSU00161543). All participants provided informed consent. Individuals were excluded if they had a history of SARS-CoV-2 infection, received other COVID-19 vaccines, or received immunosuppressive medications. No serious adverse effects were observed following heterologous or homologous boosting in this study.

### Pseudovirus neutralizing antibody assay

The SARS-CoV-2 pseudoviruses expressing a luciferase reporter gene were used to measure pseudovirus neutralizing antibodies. In brief, the packaging construct psPAX2 (AIDS Resource and Reagent Program), luciferase reporter plasmid pLenti-CMV Puro-Luc (Addgene) and spike protein expressing pcDNA3.1-SARS-CoV-2 SΔCT were co-transfected into HEK293T cells (ATCC CRL_3216) with lipofectamine 2000 (ThermoFisher Scientific). Pseudoviruses of SARS-CoV-2 variants were generated by using WA1/2020 strain (Wuhan/WIV04/2019, GISAID accession ID: EPI_ISL_402124), B.1.1.7 variant (Alpha, GISAID accession ID: EPI_ISL_601443), B.1.351 variant (Beta, GISAID accession ID: EPI_ISL_712096), B.1.617.2 (Delta, GISAID accession ID: EPI_ISL_2020950), or B.1.1.529 (Omicron, GISAID ID: EPI_ISL_7358094.2). The supernatants containing the pseudotype viruses were collected 48h after transfection; pseudotype viruses were purified by filtration with 0.45-μm filter. To determine the neutralization activity of human serum, HEK293T-hACE2 cells were seeded in 96-well tissue culture plates at a density of 1.75 × 10^4^ cells per well overnight. Three-fold serial dilutions of heat-inactivated serum samples were prepared and mixed with 50 μl of pseudovirus.

The mixture was incubated at 37 °C for 1 h before adding to HEK293T-hACE2 cells. After 48 h, cells were lysed in Steady-Glo Luciferase Assay (Promega) according to the manufacturer’s instructions. SARS-CoV-2 neutralization titers were defined as the sample dilution at which a 50% reduction (NT50) in relative light units was observed relative to the average of the virus control wells.

### Live virus neutralizing antibody assay

Full-length SARS-CoV-2 WA1/2020, and B.1.351 viruses were designed to express nanoluciferase (nLuc) and were recovered via reverse genetics. One day before the assay, Vero E6 USAMRID cells were plated at 20,000 cells per well in clear-bottom black-walled plates. Cells were inspected to ensure confluency on the day of assay. Serum samples were tested at a starting dilution of 1:20 and were serially diluted threefold up to nine dilution spots. Serially diluted serum samples were mixed in equal volume with diluted virus. Antibody–virus and virus-only mixtures were then incubated at 37 °C with 5% CO_2_ for 1 h. After incubation, serially diluted sera and virus-only controls were added in duplicate to the cells at 75 plaque-forming units at 37 °C with 5% CO_2_. Twenty-four hours later, the cells were lysed, and luciferase activity was measured via Nano-Glo Luciferase Assay System (Promega) according to the manufacturer specifications. Luminescence was measured by a Spectramax M3 plate reader (Molecular Devices). Virus neutralization titres were defined as the sample dilution at which a 50% reduction in RLU was observed relative to the average of the virus control wells.

### Enzyme-linked immunosorbent assay (ELISA)

SARS-CoV-2 spike receptor-binding domain (RBD)-specific binding antibodies in serum were assessed by ELISA. 96-well plates were coated with 2 μg/mL of SARS-CoV-2 WA1/2020, B.1.617.2 (Delta), B.1.351 (Beta), or B.1.1.529 (Omicron) RBD protein in 1× Dulbecco phosphate-buffered saline (DPBS) and incubated at 4 °C overnight. After incubation, plates were washed once with wash buffer (0.05% Tween 20 in 1× DPBS) and blocked with 350 μL of casein block solution per well for 2 to 3 hours at room temperature. Following incubation, block solution was discarded and plates were blotted dry. Serial dilutions of heat-inactivated serum diluted in Casein block were added to wells, and plates were incubated for 1 hour at room temperature, prior to 3 more washes and a 1-hour incubation with a 1:4000 dilution of anti– human IgG horseradish peroxidase (HRP) (Invitrogen, ThermoFisher Scientific) at room temperature in the dark. Plates were washed 3 times, and 100 μL of SeraCare KPL TMB SureBlue Start solution was added to each well; plate development was halted by adding 100 μL of SeraCare KPL TMB Stop solution per well. The absorbance at 450 nm, with a reference at 650 nm, was recorded with a VersaMax microplate reader (Molecular Devices). For each sample, the ELISA end point titer was calculated using a 4-parameter logistic curve fit to calculate the reciprocal serum dilution that yields a corrected absorbance value (450 nm-650 nm) of 0.2. Interpolated end point titers were reported.

### Electrochemiluminescence assay (ECLA)

ECLA plates (MesoScale Discovery SARS-CoV-2 IgG Cat No: K15463U-2; Panel 13) were designed and produced with up to 10 antigen spots in each well. The antigens included WA1/2020, B.1.617.2 (delta), and B.1.351 (beta). The plates were blocked with 50 uL of Blocker A (1% BSA in distilled water) solution for at least 30 minutes at room temperature shaking at 700 rpm with a digital microplate shaker. During blocking the serum was diluted to 1:5,000 in Diluent 100. The calibrator curve was prepared by diluting the calibrator mixture from MSD 1:9 in Diluent 100 and then preparing a 7-step 4-fold dilution series plus a blank containing only Diluent 100. The plates were then washed 3 times with 150 μL of Wash Buffer (0.5% Tween in 1x PBS), blotted dry, and 50 μL of the diluted samples and calibration curve were added in duplicate to the plates and set to shake at 700 rpm at room temperature for at least 2 h. The plates were again washed 3 times and 50 μL of SULFO-Tagged anti-Human IgG detection antibody diluted to 1x in Diluent 100 was added to each well and incubated shaking at 700 rpm at room temperature for at least 1 h. Plates were then washed 3 times and 150 μL of MSD GOLD Read Buffer B was added to each well and the plates were read immediately after on a MESO QuickPlex SQ 120 machine. MSD titers for each sample was reported as Relative Light Units (RLU) which were calculated using the calibrator.

### Intracellular cytokine staining (ICS) assay

CD4+ and CD8+ T cell responses were quantitated by pooled peptide-stimulated intracellular cytokine staining (ICS) assays. Peptide pools contained 15 amino acid peptides overlapping by 11 amino acids spanning the SARS-CoV-2 WA1/2020 or B.1.1.529 (Omicron) Spike proteins (21^st^ Century Biochemicals). 10^6^ peripheral blood mononuclear cells well were re-suspended in 100 µL of R10 media supplemented with CD49d monoclonal antibody (1 µg/mL) and CD28 monoclonal antibody (1 µg/mL). Each sample was assessed with mock (100 µL of R10 plus 0.5% DMSO; background control), peptides (2 µg/mL), and/or 10 pg/mL phorbol myristate acetate (PMA) and 1 µg/mL ionomycin (Sigma-Aldrich) (100µL; positive control) and incubated at 37°C for 1 h. After incubation, 0.25 µL of GolgiStop and 0.25 µL of GolgiPlug in 50 µL of R10 was added to each well and incubated at 37°C for 8 h and then held at 4°C overnight. The next day, the cells were washed twice with DPBS, stained with aqua live/dead dye for 10 mins and then stained with predetermined titers of monoclonal antibodies against CD279 (clone EH12.1, BB700), CD4 (clone L200, BV711), CD27 (clone M-T271, BUV563), CD8 (clone SK1, BUV805), CD45RA (clone 5H9, APC H7) for 30 min. Cells were then washed twice with 2% FBS/DPBS buffer and incubated for 15 min with 200 µL of BD CytoFix/CytoPerm Fixation/Permeabilization solution. Cells were washed twice with 1X Perm Wash buffer (BD Perm/WashTM Buffer 10X in the CytoFix/CytoPerm Fixation/ Permeabilization kit diluted with MilliQ water and passed through 0.22µm filter) and stained with intracellularly with monoclonal antibodies against IFN-γ (clone B27; BUV395), and CD3 (clone SP34.2, Alexa 700), for 30 min. Cells were washed twice with 1X Perm Wash buffer and fixed with 250µL of freshly prepared 1.5% formaldehyde. Fixed cells were transferred to 96-well round bottom plate and analyzed by BD FACSymphony(tm) system. Data were analyzed using FlowJo v9.9.

### RBD-specific B cell staining

PBMCs were stained with Aqua live/dead dye for 20 min, washed with 2% FBS/DPBS buffer, and cells were suspended in 2% FBS/DPBS buffer with Fc Block (BD) for 10 min, followed by staining with monoclonal antibodies against CD45 (clone HI30, BV786), CD3 (clone UCHT1, APC-R700), CD16 (clone 3G8, BUV496), CD14 (clone M5E2, BV605), CD19 (clone SJ25C, BUV615), CD20 (clone 2H7, PE-Cy5), IgM (clone G20-127, BUV395), IgD (clone IA6-2, PE), CD95 (clone DX2, BV711), CD27 (clone M-T271, BUV563), CD21 (clone B-ly4, PE-CF594), CD38 (clone HB7, BUV805), CD71 (clone M-A712, BV750) and staining with SARS-CoV-2 antigens including biotinylated SARS-CoV-2 (WA1/2020) RBD proteins (Sino Biological), SARS-CoV-2 (WA1/2020) RBD proteins (Sino Biological) labeled with FITC, at 4 °C for 30 min. After staining, cells were washed twice with 2% FBS/DPBS buffer, followed by incubation with BV650 streptavidin (BD Pharmingen) for 10 min, then washed twice with 2% FBS/DPBS buffer. After staining, cells were washed and fixed by 2% paraformaldehyde. All data were acquired on a BD FACSymphony flow cytometer. Subsequent analyses were performed using FlowJo software (BD Bioscience, v.9.9.6). For analyses, in singlet gate, dead cells were excluded by Aqua dye and CD45 was used as a positive inclusion gate for all leukocytes. Within class-switched B cell population gated as CD19+ IgM-IgD-CD3-CD14-CD16-, SARS-CoV-2 RBD-specific B cells were identified as double positive for SARS-CoV-2 (WA1/2020) RBD proteins labeled with different fluorescent probes. The SARS-CoV-2-specific B cells were further distinguished according to CD21/CD27 phenotype distribution.

### Statistical analysis

Descriptive statistics were calculated using GraphPad Prism 8.4.3, (GraphPad Software, San Diego, California). Data are presented as medians with interquartile ranges (IQR).

**Table S1.**
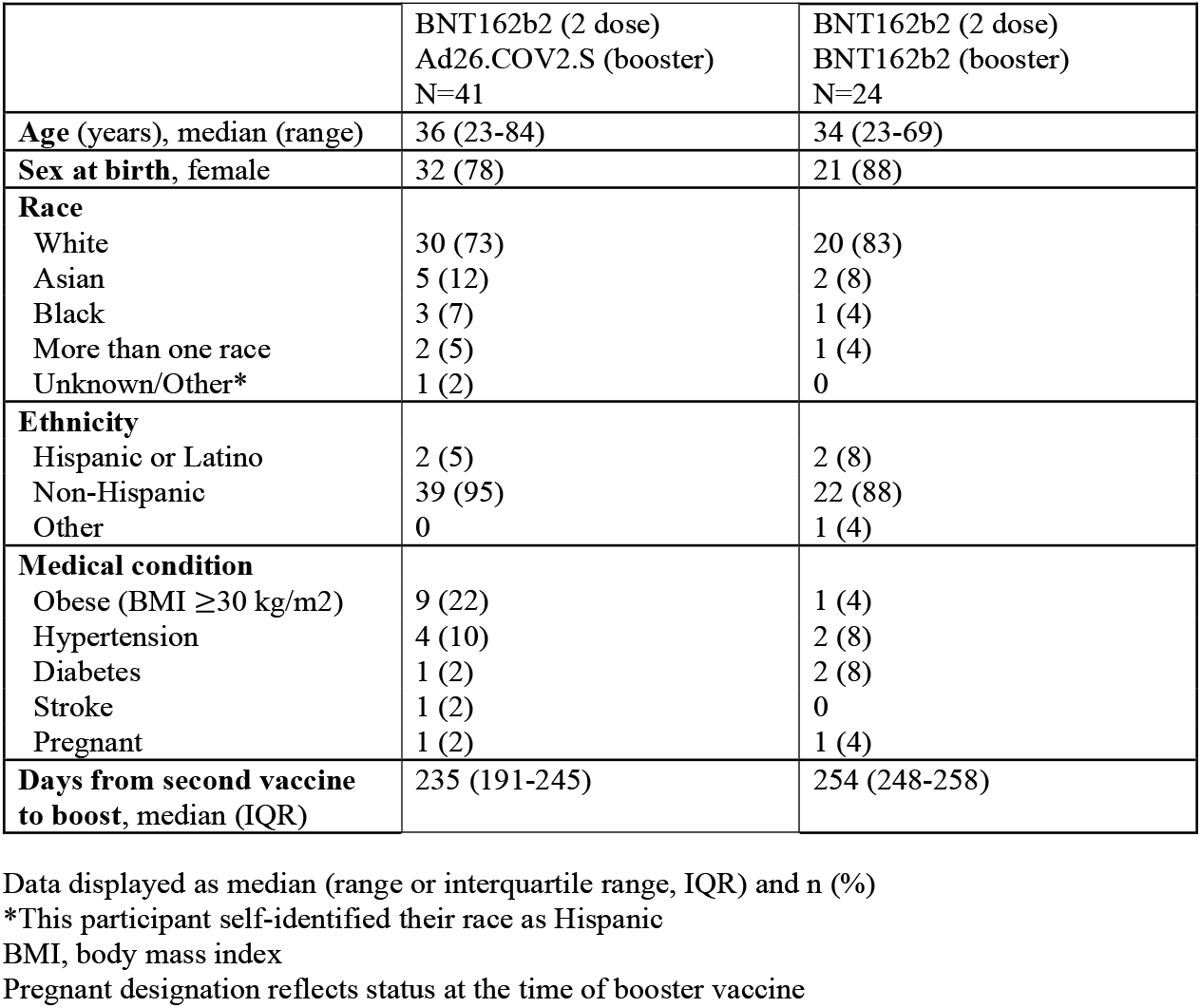
Characteristics of the study population

**Table S2.** Ratio of NAb titers (week 4 / week 2)

|  | **Ad26.COV2.S Boost** |  |  | **BNT162b2 Boost** |  |  |
| --- | --- | --- | --- | --- | --- | --- |
|  | **Week 2** | **Week 4** | **Ratio** | **Week 2** | **Week 4** | **Ratio** |
| **WA1/2020** | 1,462 | 3,597 | 2.46 | 7,554 | 3,986 | 0.53 |
| **Delta** | 1,009 | 2,148 | 2.13 | 3,040 | 1,541 | 0.51 |
| **Beta** | 899 | 1,924 | 2.14 | 1,865 | 1,472 | 0.79 |
| **Omicron** | 591 | 859 | 1.45 | 1,018 | 345 | 0.34 |

## Supplementary Figure Legends

**Figure S1.**
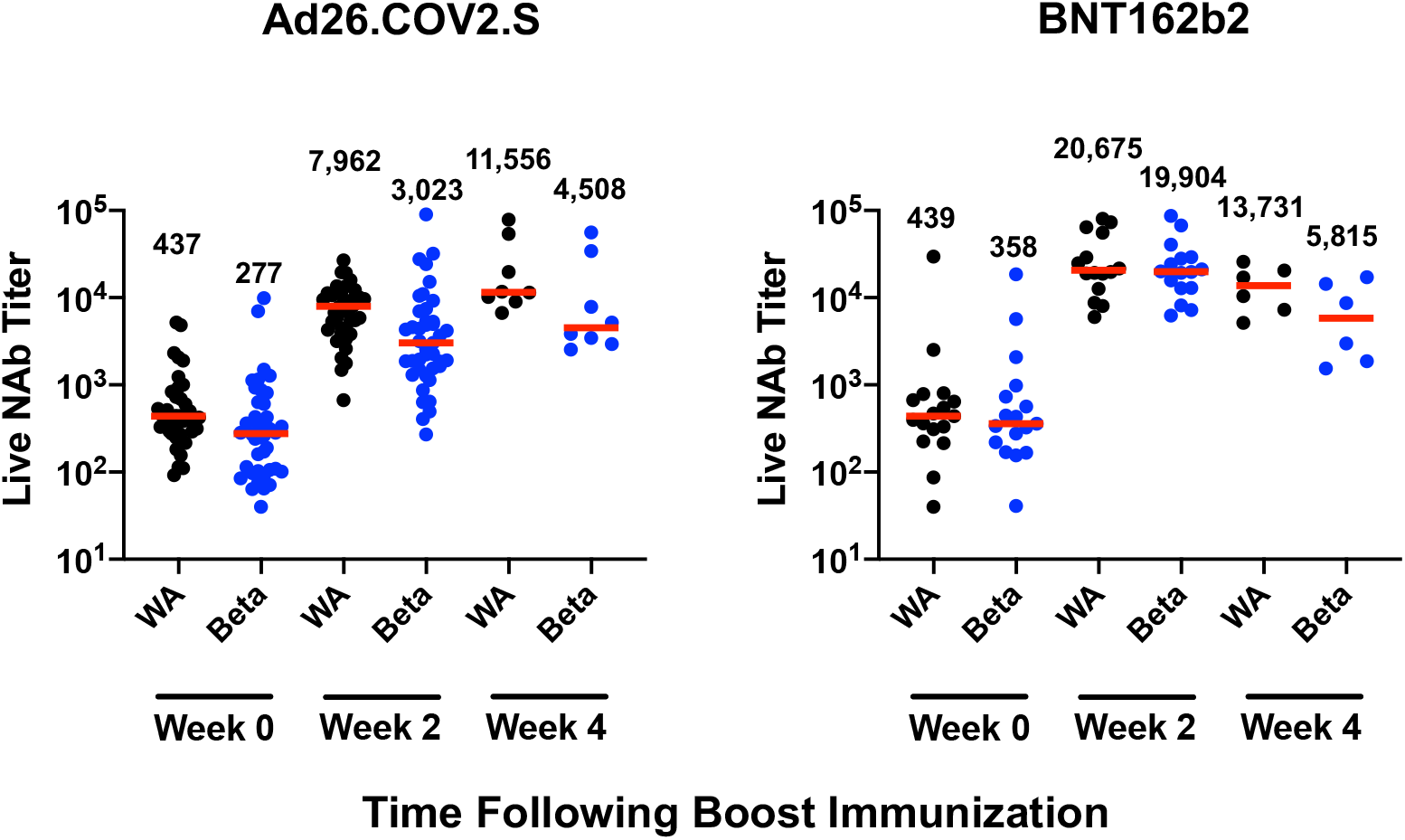
Live virus neutralizing antibody responses following Ad26.COV2.S and BNT162b2 boosting. Live virus neutralizing antibody titers at weeks 0, 2, and 4 following boosting BNT162b2 vaccinated individuals with Ad26.COV2.S or BNT162b2 were assessed against the SARS-CoV-2 WA1/2020 and B.1.351 (Beta) variants. Medians (red bars) are depicted and numerically shown. A subset of samples were available at week 4.

**Figure S2.**
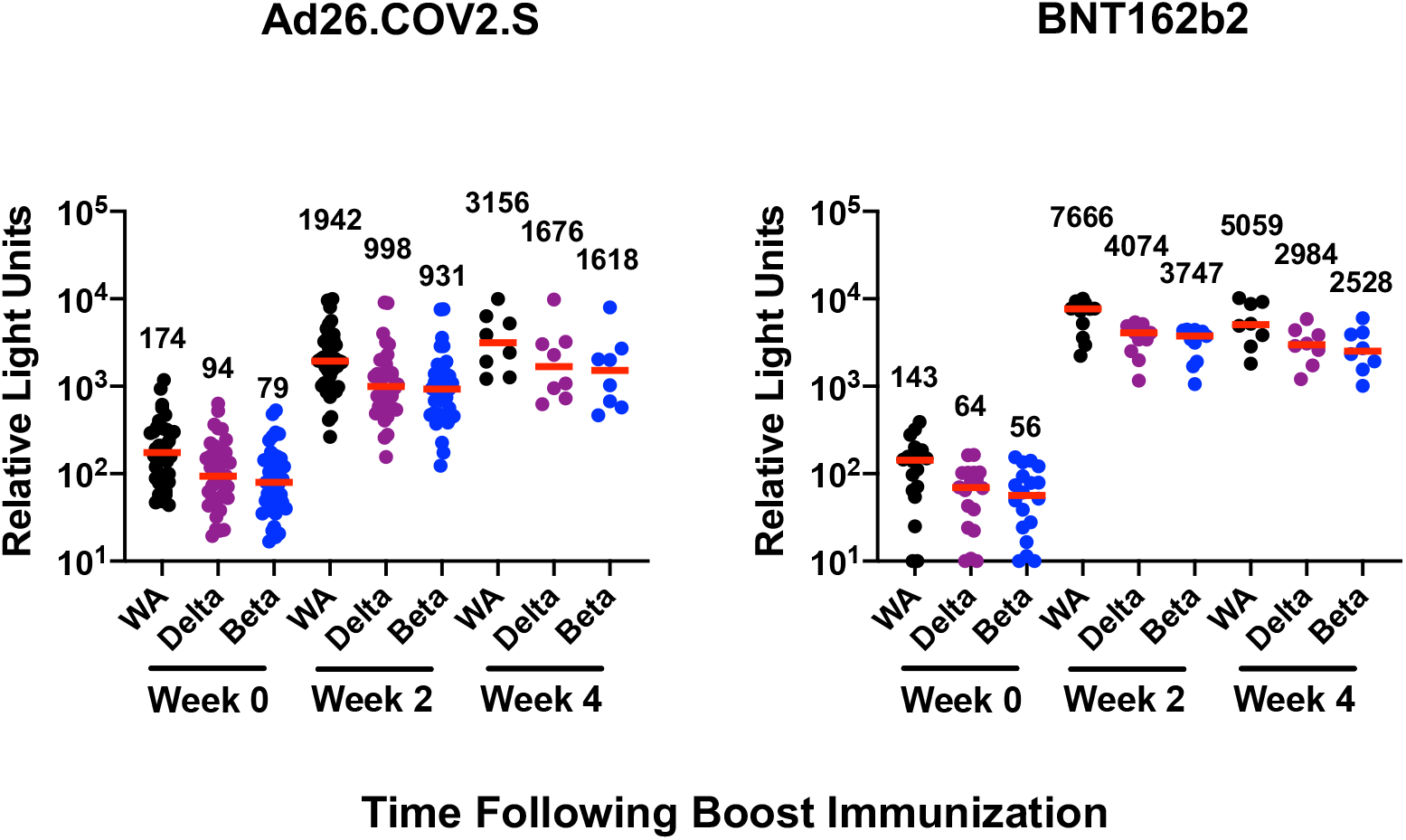
ECLA binding antibody responses following Ad26.COV2.S and BNT162b2 boosting. Binding antibody titers at weeks 0, 2, and 4 following boosting BNT162b2 vaccinated individuals with Ad26.COV2.S or BNT162b2 were assessed by electrochemiluminescence binding antibody assay (ECLA) against the SARS-CoV-2 WA1/2020, B.1.617.2 (Delta), and B.1.351 (Beta) strains. Medians (red bars) are depicted and numerically shown. A subset of samples were available at week 4.

**Figure S3.**
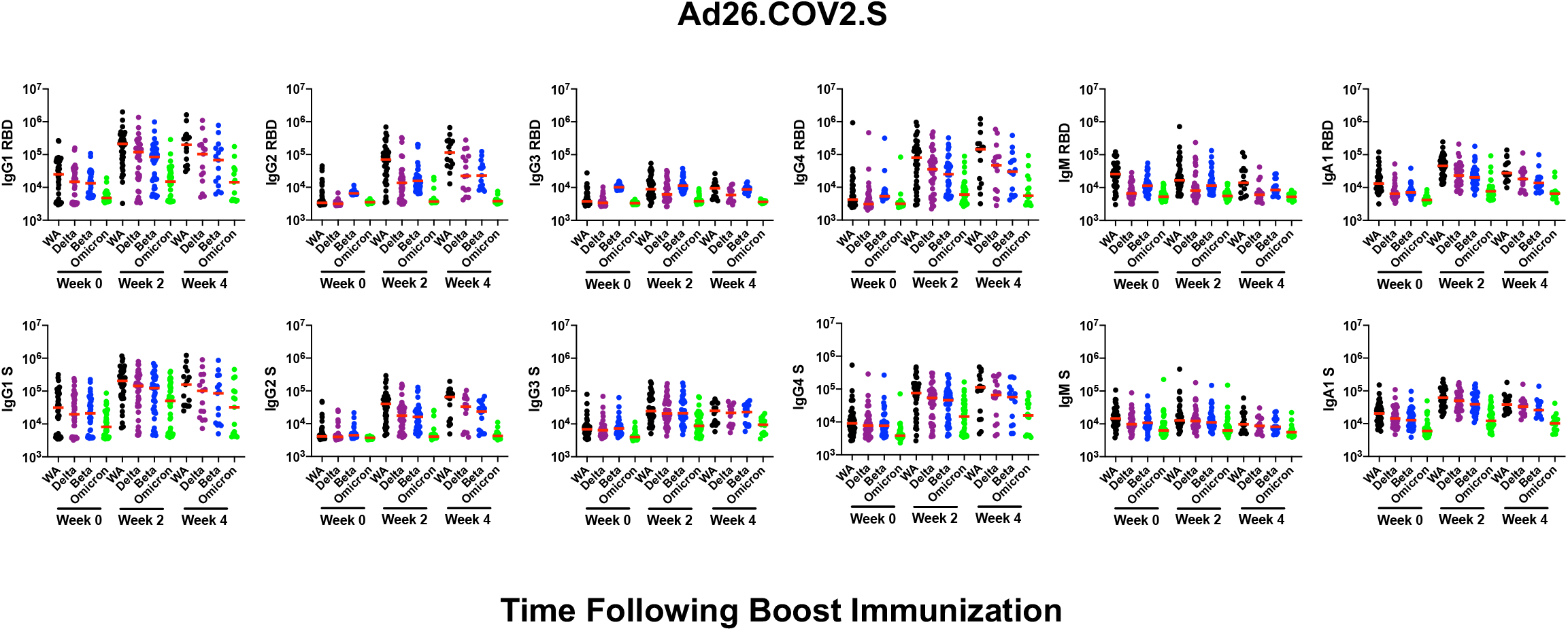
Isotypes of antibody responses following Ad26.COV2.S and BNT162b2 boosting. IgG1, IgG2, IgG3, IgG4, IgM, and IgGA1 antibody titers at weeks 0, 2, and 4 following boosting BNT162b2 vaccinated individuals with Ad26.COV2.S against the SARS-CoV-2 WA1/2020, B.1.617.2 (Delta), B.1.351 (Beta), and B.1.1.529 (Omicron) strains. Medians (red bars) are depicted and numerically shown. A subset of samples were available at week 4.

**Figure S4.**
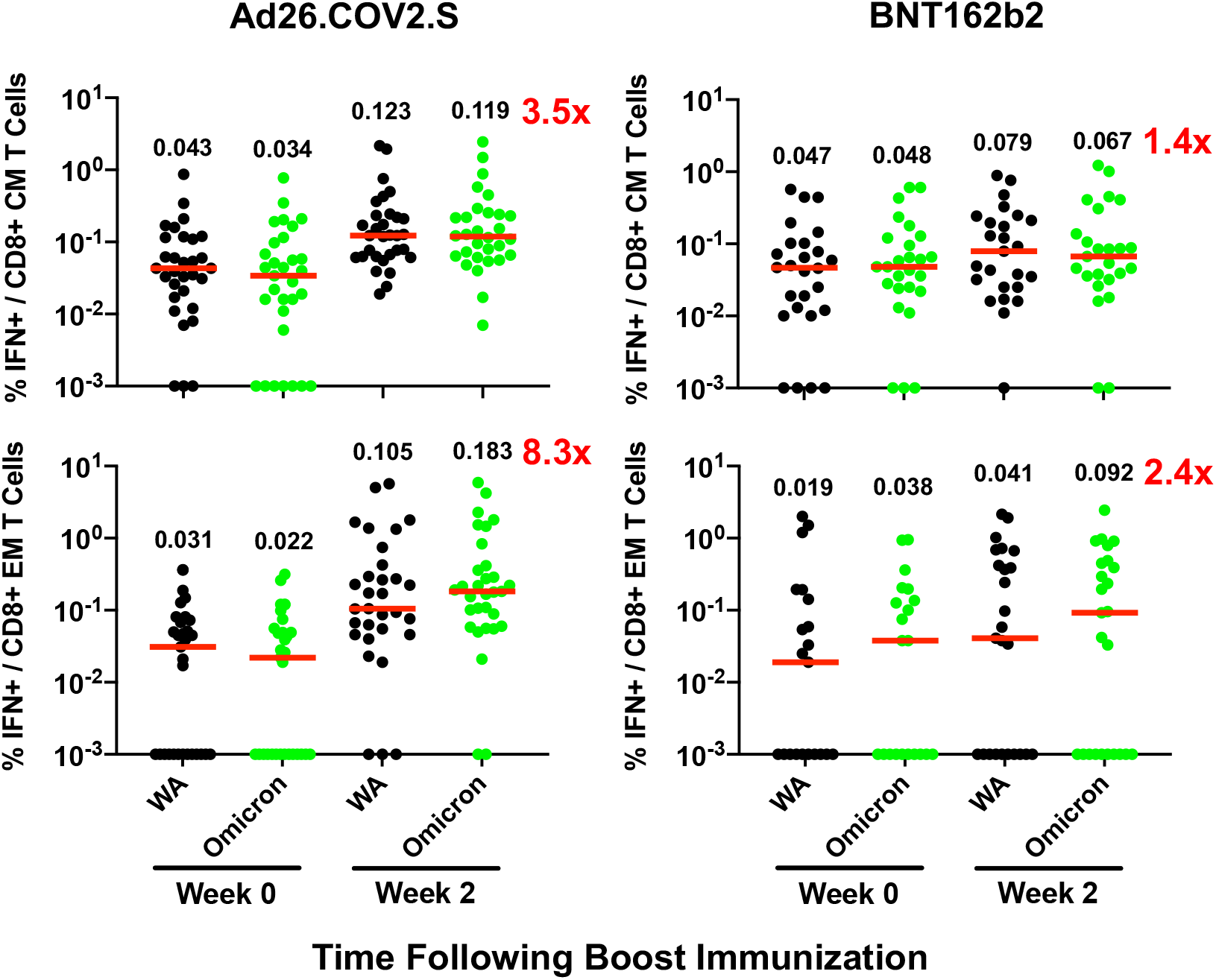
CD8+ T cell memory responses following Ad26.COV2.S and BNT162b2 boosting. Pooled peptide IFN-*γ* CD8+ central memory (CM) and effector memory (EM) T cell responses by intracellular cytokine staining assays against the SARS-CoV-2 WA1/2020 and B.1.1.529 (Omicron) variants. Medians (red bars) are depicted and numerically shown.

**Figure S5.**
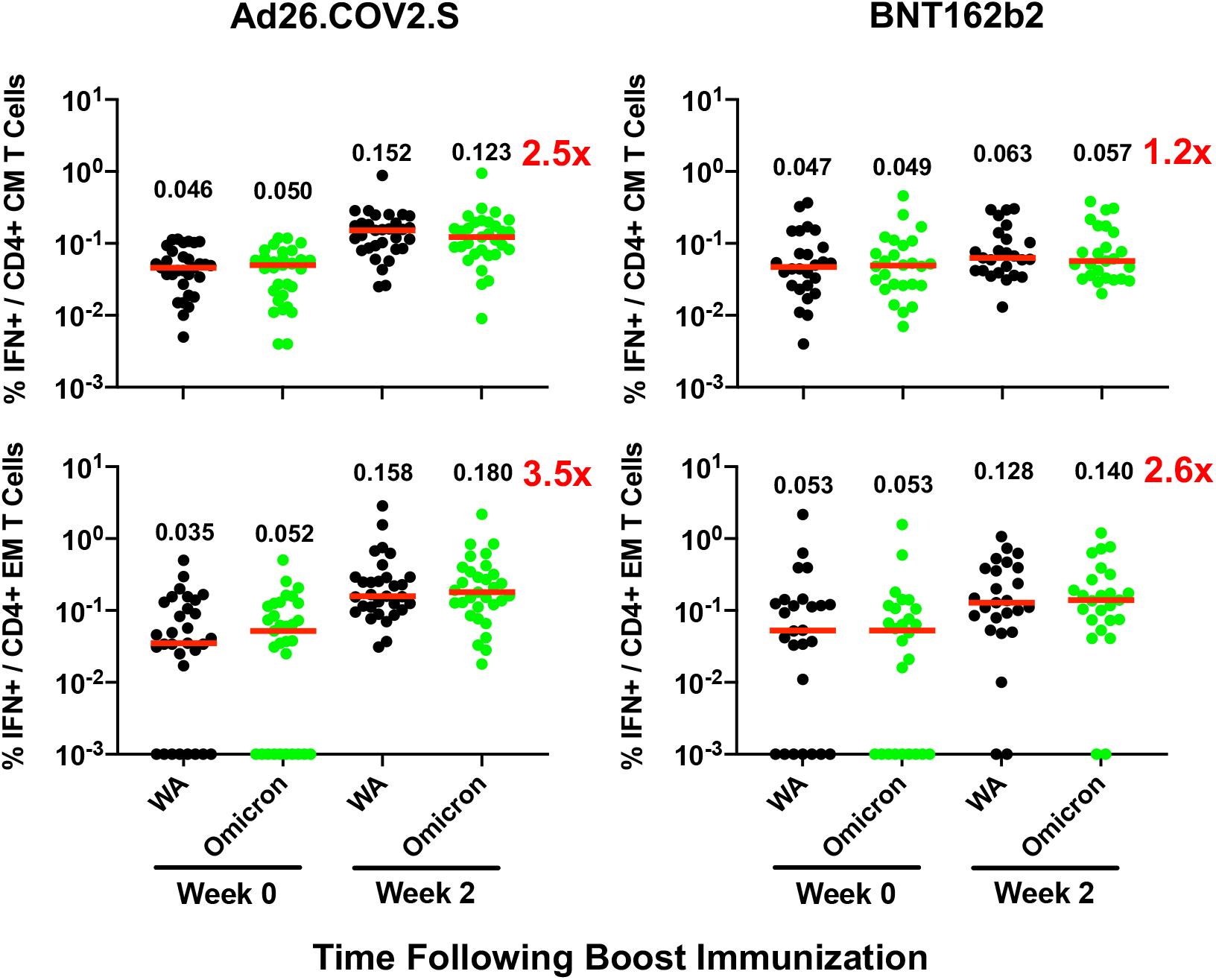
CD4+ T cell memory responses following Ad26.COV2.S and BNT162b2 boosting. Pooled peptide IFN-*γ* CD4+ central memory (CM) and effector memory (EM) T cell responses by intracellular cytokine staining assays against the SARS-CoV-2 WA1/2020 and B.1.1.529 (Omicron) variants. Medians (red bars) are depicted and numerically shown.

**Figure S6.**
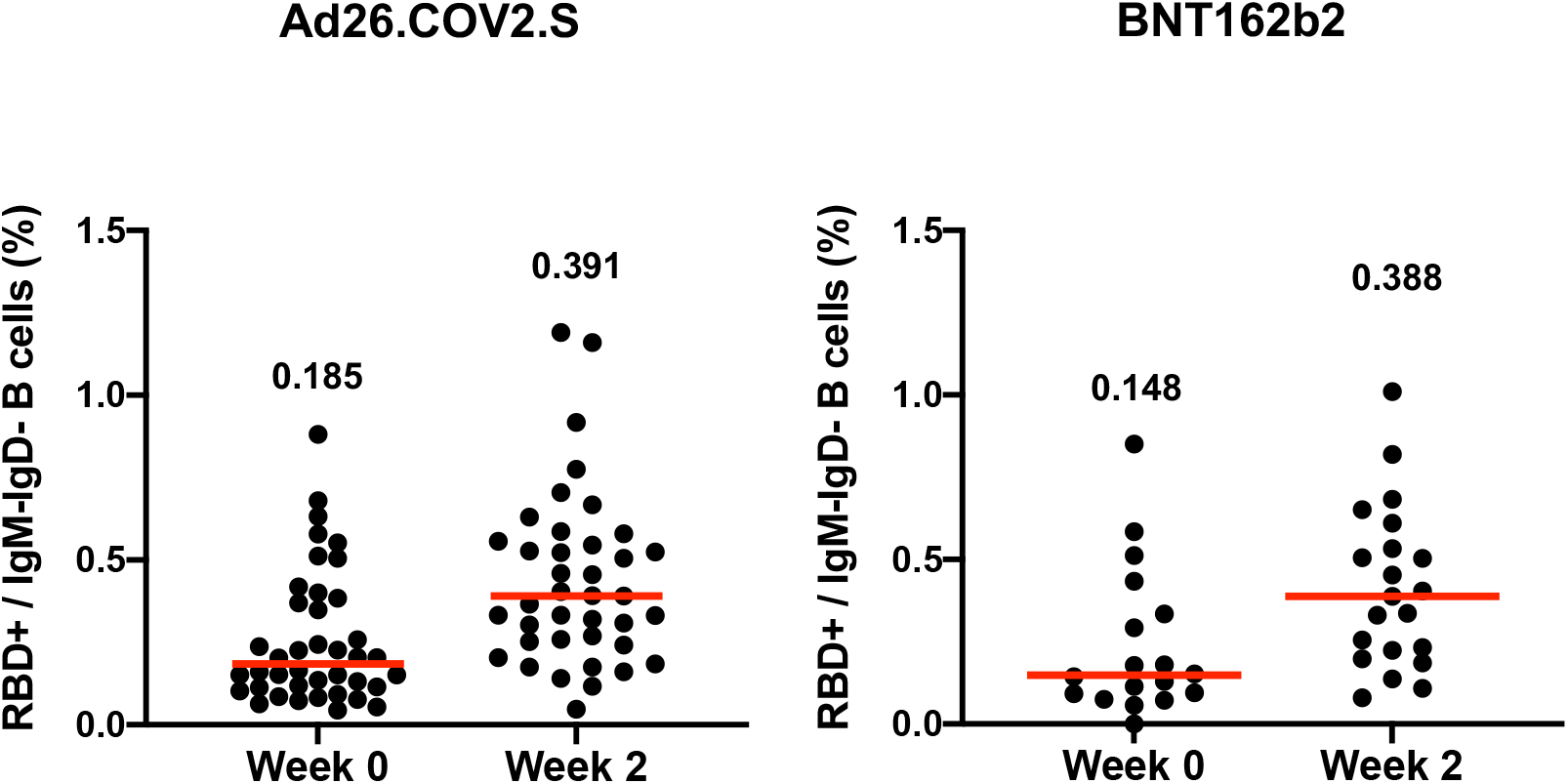
Levels of WA1/2020 RBD-specific memory B cells. Receptor binding domain (RBD)-specific memory B cells at weeks 0 and 2 following boosting BNT162b2 vaccinated individuals with Ad26.COV2.S or BNT162b2. Medians (red bars) are depicted and numerically shown.

**Figure S7.**
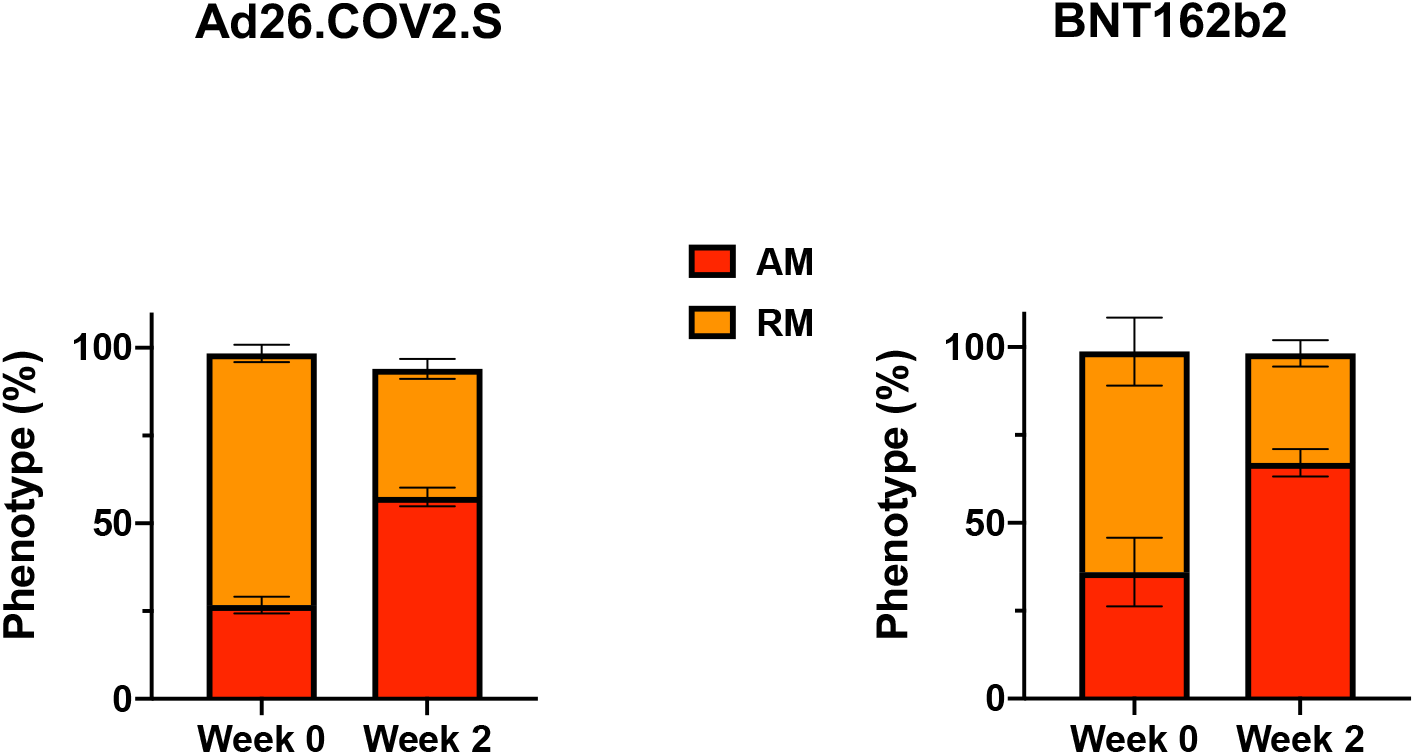
Phenotype of WA1/2020 RBD-specific memory B cells. Phenotype of RBD-specific memory B cells at weeks 0 and 2 following boosting BNT162b2 vaccinated individuals with Ad26.COV2.S or BNT162b2. Red, activated memory (AM) B cells. Orange, resting memory (RM) B cells.

**Figure S8.**
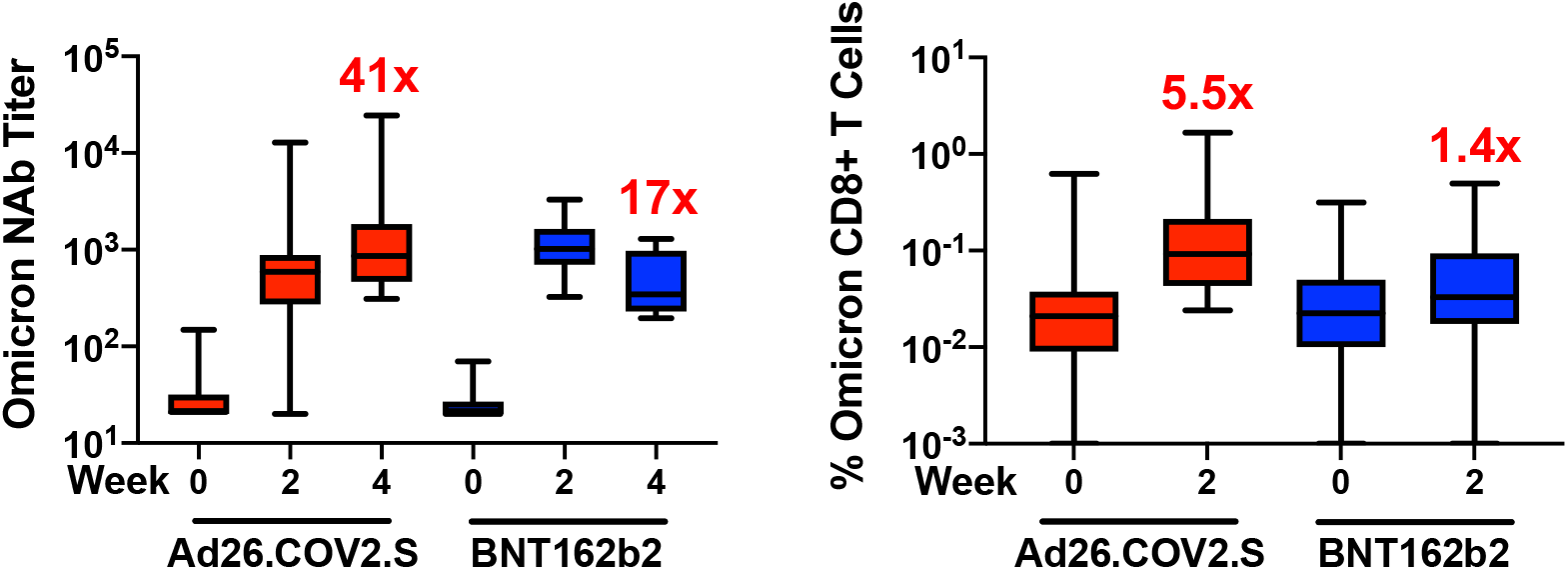
Summary of Omicron-specific neutralizing antibody and CD8+ T cell responses following Ad26.COV2.S and BNT162b2 boosting. Box-and-whisker plots are shown. Numbers reflect fold increase following the boost compared with prior to the boost.

